# A study protocol for the project *Meeqqat Peqqissut:* A register-based study exploring social and health determinants of child well-being in Kalaallit Nunaat (Greenland)

**DOI:** 10.64898/2026.06.25.26356634

**Authors:** Charlotte Brandstrup Ottendahl, Ivalu Katajavaara Seidler, Lars Pedersen, Anders Blaabjerg, Christina Viskum Lytken Larsen

**Author notes:** Corresponding Author: Charlotte Brandstrup Ottendahl, Studiestrædet 6, 1455 Copenhagen.

## Abstract

**Introduction:** Child health and well-being in Kalaallit Nunaat (Greenland) remain a major public health concern, yet systematic, population-level research is limited. Previous studies have focused on isolated indicators such as vaccination coverage and infant mortality, without capturing the broader structural and social determinants shaping children’s lives. The Meeqqat Peqqissut project addresses this gap by developing a cross-sectoral, register-based approach to identify both risk and health promoting factors influencing child health and well-being for alle children in Kalaallit Nunaat. Grounded in the Indigenist Ecological Systems Model (IESM) and complemented by the Peqqissuserput framework, the project adopts a strengths-based perspective that emphasizes resilience and culturally embedded determinants of well-being.

**Methods:** Meeqqat Peqqissut is a collaboration between the Centre for Public Health in Greenland and Statistics Greenland. The study integrates longitudinal register data linked via personal identification numbers, combining healthcare records (COSMIC), socio-economic data from Statistics Greenland, municipal case management systems (Get Organized), and police records. Additional sources include education and social services. Data will be combined and analysed on Statistics Greenland’s secure platform using advanced methods such as latent class and trajectory analyses. A participatory approach ensures continuous dialogue with practitioners to validate data quality and contextual relevance.

**Results:** The project supports evidence-based strategies for improving child health and well-being in Kalaallit Nunaat and contributes to the national public health strategy Inuuneritta III. It aligns with the Kalaallit Nunaat government’s research strategy for 2022–2030, as it aims to support sustainable societal development by contributing to improved public health and reducing social health inequalities. Ethical approval has been obtained from the Scientific Ethics Committee in Kalaallit Nunaat (VEK 2025-12).

**Conclusion:** The *Meeqqat Peqqissut* project will provide a robust, cross-sectoral evidence base to strengthen efforts to improve child health and reduce social health inequalities in Kalaallit Nunaat. By integrating administrative data and applying an Indigenous, strengths-based framework, the study advances understanding of structural and social determinants of well-being. The project establishes a sustainable approach to equity-oriented child health monitoring and will inform policy, practice, and long-term public health action.

## Background

The health and well-being of children in Kalaallit Nunaat are widely debated, yet systematic research on their overall health status and the key social determinants influencing their well-being remains limited. While previous studies have examined specific health outcomes, a comprehensive, population-level understanding of children’s health and equities herein has not been prioritized. This study addresses this gap by introducing a novel, cross-sectoral research approach that both identifies risk factors and health promoting factors influencing well-being and health outcomes among all children I Kalaallit Nunaat.

Despite years of research on children’s health in Kalaallit Nunaat, efforts have been fragmented. In their efforts to establish national indicators for children’s health, Niclasen and Köhler concluded that a lack of both validated data sources and routine data collections were major obstacles for systematic monitoring of child health (1). While some studies have utilized national register data, they have primarily focused on isolated health indicators such as vaccine coverage, dental status, infant mortality, acute otitis media, heart disease and weight and have not supported systematic monitoring in Kalaallit Nunaat (2–8). Additionally, past cohort studies have contributed valuable insights into prenatal exposures, disease patterns, and anthropometric measures etc. but have limited national coverage (9–13).

Children across Kalaallit Nunaat are exposed to multiple health risk factors. Cross-sectional data from the latest Population Health Survey in Kalaallit Nunaat (2018) indicates that approximately a third of youth (15-34 years) have experienced adverse childhood experiences (ACEs), such as alcohol problems, violence, and sexual abuse, before the age of 18. These early-life ACEs significantly increase the risk of negative health outcomes throughout life (14–16). The impact of these challenges is reflected in persistently high youth suicide rates, placing considerable strain on the healthcare system and society as a whole (17, 18).

In Kalaallit Nunaat, the data infrastructure for continuous public health monitoring through registers has not been prioritized. Despite growing political awareness regarding child health issues, the absence of comprehensive, population-level research continues to hinder the development of evidence-based preventive strategies tailored to the context (19). Valid longitudinal data are essential for enabling early interventions that prevent adverse childhood experiences (ACEs) from escalating. Such data support proactive strategies to identify and mitigate risks during early development, reducing reliance on secondary prevention that primarily addresses consequences. Until recently, the lack of routinely collected and validated data on child health severely restricted register-based and longitudinal research (1). However, recent advancements in electronic registration systems offer new possibilities for advanced epidemiological research on longitudinal data across sectors. In other Indigenous Arctic settings, longitudinal studies such as the SamBa study and the Arctic Childhood Study aim to provide systematic data for monitoring vulnerable Sami children in Norway (20, 21). Similar approaches in larger Nordic societies, like Denmark, include the Child Health Database, which has enabled health profiles for birth cohorts since 2002 (22) and supports municipal preventive work, with a clinical database under development (23, 24). Likewise, the establishment of the Danish DANLIFE cohort has enabled new research on how ACEs affect health across the life course (25).

This study aims to leverage recent advancements in Kalaallit Nunaat to investigate the key determinants of child health by integrating register data across sectors. It is grounded in a strength-based approach, highlighting both risk and health promoting factors, with particular attention to the resources that support children’s resilience and holistic well-being. This protocol outlines the procedures for the research project Meeqqat Peqqissut (Healthy Children in Kalaallit Nunaat).

## Theoretical Frame

Meeqqat Peqqissut is based on two theoretical approaches:

### The Indigenist Ecological Systems Model

The theoretical foundation for Meeqqat Peqqissut draws upon the social determinants of health (SDH) framework, which emphasizes that health outcomes are influenced not only by individual factors but also by the broader social, economic, and environmental conditions in which individuals live (26, 27). This perspective is integrated within the Indigenist Ecological Systems Model (IESM), developed by Fish, Hirsch and Syed (28) and exemplified by O’Keefe, Fish (29), which extends Bronfenbrenner’s ecological systems theory to better account for the unique historical, cultural, and environmental contexts of Indigenous Peoples. While Bronfenbrenner’s original model outlines the interaction between nested systems, from the immediate microsystem to broader societal structures (30), the IESM reconfigures these systems to prioritize the role of historical trauma, colonization, and cultural traditions in shaping Indigenous health and well-being (29).

In this way, the IESM represents an ontological standpoint that recognizes the interconnectedness of individual, familial, cultural, ecological, and historical influences on health, offering a more holistic and culturally relevant framework for understanding health in Indigenous contexts (28). This model is particularly important in Kalaallit Nunaat, where child health is shaped not only by familial and community factors but also by broader social, historical, and ecological environments, including the legacies of colonization and the enduring significance of Inuit culture. IESM promotes a strengths-based approach by focusing on the resilience, cultural knowledge, and community strengths that contribute to Indigenous health, rather than deficits (Baskin, 2022; Kirmayer, Dandeneau, Marshall, Phillips, & Williamson, 2011). This aligns with Cueva et al. (2021), who emphasize how research framework in health should be community-driven and strength-based and the cultural and ecological contexts.

### A theoretical research model starting from a place of strengths to support Kalaallit Inuit communities to thrive

The Peqqissuserput framework offers a holistic view of health, ethical principles for research, and exemplifies determinants of health and well-being in the context of Kalaallit Nunaat, seeing it as a dynamic state influenced by several aspects that are interconnected (31). This framework complements the IESM, emphasizing the interconnectedness of social, cultural, environmental, and ecological factors in shaping health and well-being, and underlines the importance of strengths-based, community-driven research. Furthermore, the Meeqqat Peqqissut project adheres to the ethical principles outlined in the Peqqissuserput framework, ensuring that all research activities are carried out with respect for SILA and are guided by these holistic and culturally grounded values.

## Methods

### Study Design

The research project is a collaboration between The Centre for Public Health in Greenland and Statistics Greenland. The project includes a PhD project and a postdoctoral project. Both the PhD and postdoc projects are designed as longitudinal register-based studies, incorporating nationwide data of children up to the age of 18 as study population (32, 33). The PhD project will primarily use healthcare system data to investigate how socioeconomical status in the first 1000 days of life impact early childhood health. The postdoctoral project will extend the research to children aged 2-18 years, combining data from various sectors such as healthcare, municipalities, and the police to form the basis of explorative analysis into current and later development of child health.

### Data sources and management

All data will be integrated an analysed through Statistics Greenland’s secure platform, data will include registrations for all children until age 18 including prenatal care registrations. On average app. 750 children are born every year in Kalaallit Nunaat (34). Based on results from a preceding pilot study we expect that data from at least 2020 and going forward are of high quality, but children will be included retrospectively as far back as registrations allow resulting in an estimated study population of at least 4400+ children (34) . The child’s personal identification number (CPR) will be used to link data from various sources, enabling the tracking of children’s health and social determinants over time (35). Additionally, parental data, such as employment, education and income, will be linked to children’s data through Statistics Greenland’s registries.

We place strong emphasis on collecting recent and prospective data, as historical data can be difficult to validate. While historical data from Statistics Greenland will be tested and utilized our focus is to build a robust longitudinal dataset. Comprehensive metadata will be created ensuring continuous updates on the data and accessibility for others, including future users of Statistics Greenland’s research portal. Figure 1 provides an overview of the available data sources to be included in the project, along with a brief presentation of additional potential data sources. The different data sources will be further elaborated in the following.

1. **Data from the health care system.** Data from the health care system is stored in the electronic patient journal (EPJ) system COSMIC. COSMIC is widely used in various healthcare settings, including university hospitals, primary care clinics etc. COSMIC offers cohesive and patient-focused operational support, ensuring that essential patient information is readily accessible to healthcare professionals (36). In Kalaallit Nunaat the health care system includes hospitals (including prenatal care, births, acute treatment, physical therapy, other hospitalizations), primary health care, children’s dental care and community health nurses. COSMIC was implemented in the healthcare system between 2013 and 2016 (18), but full implementation was achieved in some places in 2019. The implementation did not include the Eastern town Tasiilaq, where the previously used EPJ-system ÆSKULAP is still in use due to technical difficulties. ÆSKULAP was implemented in 2007, except for most units at Queen Ingrids hospital in Nuuk, where paper journals were used until the implementation of COSMIC. Before ÆSKULAP the community health nurses used the EPJ system NOVAX. Plans are underway to replace COSMIC, as the developer will stop supporting the system in a foreseeable future (18). Although a replacement system has not yet been selected, the process has begun (37). Hospital data and primary care data from COSMIC have previously been used for scientific research (2–8). Data from community health nurses has also been used but to little extent (12). To assess the validity of data in the health care system (especially data from the community health nurses) for research purposes, the authors conducted a pilot study involving expert interviews with midwifes, community health nurses and doctors (38). The study yielded important knowledge on registration practices within the health care system and on the potential use of these data for monitoring purposes (39).
2. **Data from Statistics Greenland**. Statistics Greenland provide data on socio-economic information on the population in Kalaallit Nunaat, on individual, family, and community levels. Data includes parents’ age, place of residence, income, education, and employment status. Additionally, Statistics Greenland hosts the Birth Register, which covers the period from 1989–2018 and are registered manually from birth certificates. Since 2019, data has been electronically imported from COSMIC. Statistics Greenland primarily compiles data from administrative systems, which means that they have data on education and social conditions. This comprehensive data is used not only for political decision-making and public service planning but also for research.
3. **Municipal data.** Municipalities in Kalaallit Nunaat registers data on children’s health and well-being as part of their social service efforts. However, this work was significantly strengthened in 2021 with the launch of the Kalaallit Nunaat-Danish cross-sectional collaboration. This was a collaboration between Kalaallit Nunaat and Danish authorities aiming at enhancing social services for children and youth. The partnership involved the five municipalities in Kalaallit Nunaat, social services in both countries, the Agency of Education and Quality in Denmark, the Ministry of Social Affairs and Senior Citizens, the Ministry of Justice, and Kalaallit Nunaat’s Department for Children, Youth, and Families. One of the pivotal aspects of this collaboration was Initiative 6, which focuses on the implementation of standardized case management procedures across all five municipalities, and assisting municipalities in adopting a new, unified IT system called Get Organized for case management (40). Since January 2024, it has been mandatory for municipalities to conduct case management in accordance with the Inatsisartut Act on Support for Children using a shared public IT system (41). For the municipalities’ new ESDH system, Get Organized, workflows have been developed for the 14 most common case types, including notifications, child welfare assessments, and out of home placements. Step-by-step guidelines have been created for all workflows, and the Kalaallit Nunaat Agency of Social Affairs has also developed and quality-assured templates that have been integrated into the system. Consequently, all municipalities are now required to use the same case management procedures and templates, and using the same guides (42). The introduction of these standardized case management practices and the Get Organized IT system is a substantial step towards improving the integrity, reliability, and usability of data in the child welfare system for research in Kalaallit Nunaat.
4. **Data from the police in Kalaallit Nunaat.** The police maintain a database containing information related to law enforcement activities across the country. This includes reports on criminal activities, investigations, arrests, and other actions taken by law enforcement. As a new initiative the police also register when alcohol is involved in an episode requiring their assistance. The data is used by the police for operational purposes, including tracking ongoing investigations, managing cases, and ensuring effective law enforcement across municipalities. Additionally, the police publish annual reports, available on their official website. These reports offer valuable insights into crime trends, law enforcement efforts, and statistics on criminal activities in Kalaallit Nunaat. For this project, the police database will be used to provide data related to children’s exposure to violence, abuse, and other traumas, which is crucial for understanding and addressing the impact of these factors on children and youth.
5. **Other relevant data sources** Several other cross-sectoral data sources offer valuable insights into children’s health and well-being in Kalaallit Nunaat. The Agency of Education collects data annually on October 1st, which includes information on the number of children in special education, associated diagnoses, and academic outcomes such as exam results. The national advisory service on special education (MISI) and municipal handicap teams may also hold relevant data, including developmental assessments, school readiness tests, and records of referrals to specialist care. From the social sector, data from publicly operated residential institutions, collected by the social agency, and information on parental addiction treatment from the national treatment center Allorfik could shed light on unhealthy family environments. The justice system, including the Prison and Probation Service, may provide data on incarcerated parents, while additional health and social data may be available from Steno Diabetes Center Greenland and lifestyle clinics. Non-digital health assessments conducted in daycare settings may offer further insights into early child development. The free help line “Tusaannga” also has data on children and youth who call the help service. Finally, local sports associations may hold records on children’s participation in organized activities, offering rare indicators of health promoting factors and community engagement.

**Figure 1:**
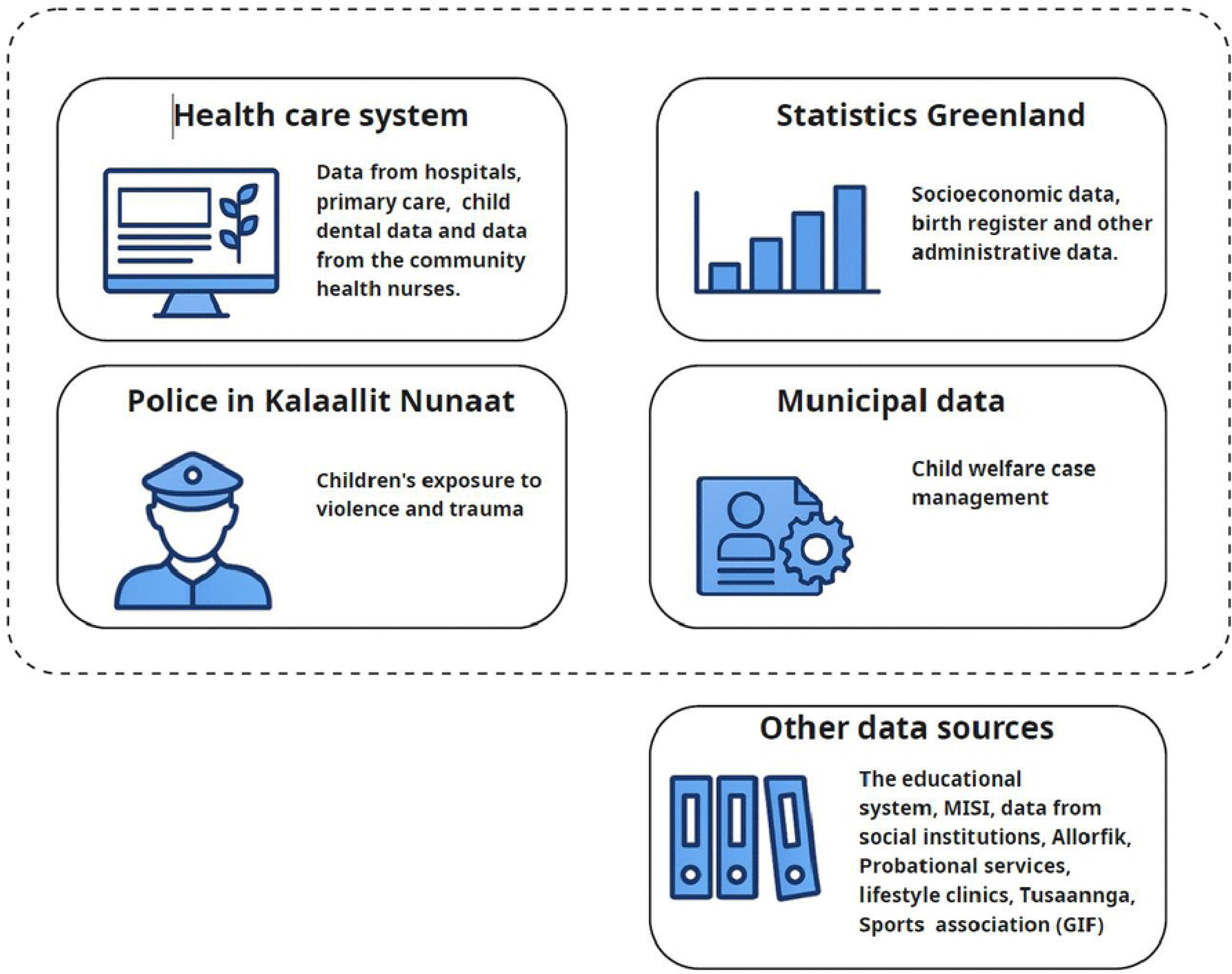
Overview of available data sources for analyses. Figure generated with Microsoft co-pilot.

### Patient and Public Involvement - Participatory approach to data validation and management

Ensuring valid and contextually meaningful data requires more than technical infrastructure, it demands continuous dialogue with practitioners and other relevant knowledge holders. This approach is particularly important for understanding the conditions under which data is registered and the data quality and completeness. In line with principles of community-based participatory research (43, 44), we actively involve knowledge holders throughout the data collection, management, and analysis. Their insights contribute to incorporation of local practices and institutional realities. As part of this process, we will engage in dialogue with all five municipalities to discuss their data and registration practices in detail. Furthermore, we prioritize reporting data back to practitioners. This feedback loop supports reflection on data registration practices and promotes shared responsibility for data quality. This participatory model not only strengthens the validity of the data but also enhances local relevance and sustainability of the research process.

### Operationalizing strengths-based approaches in register research

Strengths-based approaches are increasingly recognized in health research, particularly in studies led by Indigenous scholars and Arctic research teams, for their capacity to elucidate resilience, community assets, and culturally embedded conceptions of well-being (45). These paradigms provide a critical counterpoint to prevailing deficit-oriented models by emphasizing the determinants that foster health and thriving communities, rather than focusing exclusively on risk and pathology.

Despite their promise, strengths-based perspectives have seldom been systematically operationalized in register-based research. Administrative datasets are predominantly structured to capture adverse outcomes, such as diagnoses, service utilization, or negative events, which complicates the identification and measurement of health promoting factors. In this study, we seek to address this gap by incorporating both risk and health promoting factors into our analyses, drawing on integrative frameworks such as Peqqissuserput and the IESM. These frameworks foreground holistic, relational, and contextually grounded understandings of health and well-being.

It is important to acknowledge that the identification of culturally relevant indicators and health promoting factors within register-based data is inherently exploratory. While clinical practitioners working with children and families routinely recognize strengths and resources in practice, there is limited empirical knowledge regarding the extent to which such health promoting factors are systematically recorded. To address this, we will investigate the presence and documentation of health promoting factors within relevant data sources. In the health care system free-text data registrations from the community health nurses and midwifes will be investigated. Additionally, in collaboration with municipal stakeholders, we will examine which health promoting factors are captured in their administrative systems and evaluate the feasibility of integrating these data into register-based research. Guided by the IESM framework, our approach aims to identify health promoting factors at both the individual and community levels.

### Research Plan

The project is structured into three work packages (WPs), shown in the Gantt chart in Figure 2. The three work packages will be introduced in the following.

**Figure 2:**
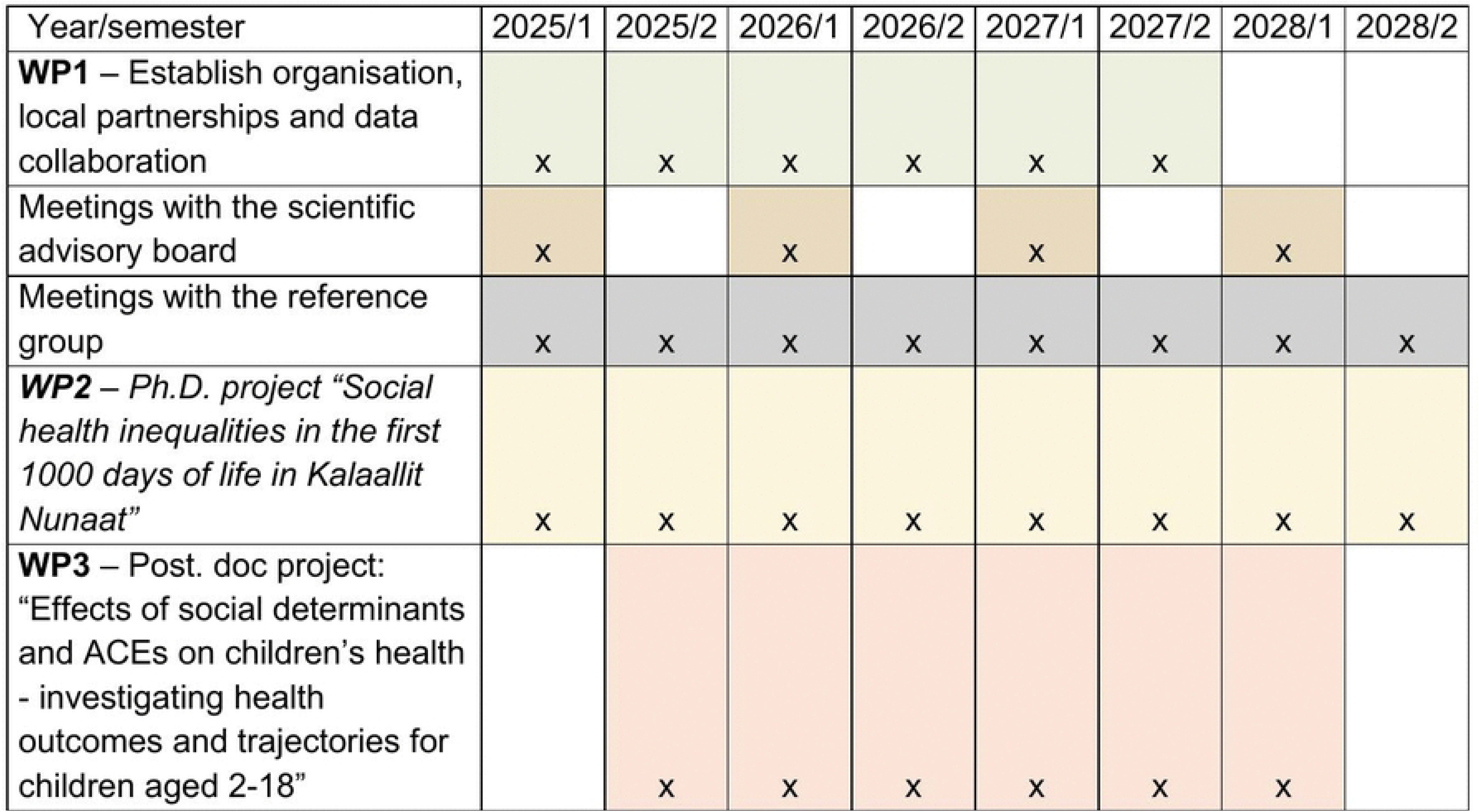
Research Plan Meeqqat Peqqissut, 2025-2028, Gannt chart

#### WP1 – Establish organisation, local partnerships and data collaboration

WP1 focuses on building the organisational and collaborative infrastructure necessary for the success of the overall project. The project group consists of the Centre for Public Health in Greenland and Statistics Greenland. This work package includes forming a scientific advisory board and a reference group, as well as developing a comprehensive data management plan for integrating datasets from multiple sectors. The two groups are formed to ensure scientific quality and practical relevance and establishing cross-sector collaboration among key data holders, such as healthcare, municipalities, social services, and the police, is a central component of this work package.

The scientific advisory board is composed of researchers with expertise in child health, epidemiology, psychology, and statistics who provide methodological guidance and quality assurance. The reference group includes professionals from a wide range of institutions working with children’s health, welfare, and social conditions in Kalaallit Nunaat, representing sectors such as healthcare, social services, education, law enforcement, and government agencies. This broad, cross-sector collaboration ensures that the project benefits from diverse perspectives and practical experience, contributing to relevant and applicable outcomes.

The scientific advisory board meets annually, while the reference group convenes twice a year, with the possibility to form smaller working groups to address specific data issues or thematic areas. WP1 also includes a comprehensive review of existing literature to inform the project’s methods and contextualize its findings. In addition, international collaborations with Arctic health research institutions strengthen the project’s knowledge base and capacity.

WP1 thus lays the essential groundwork for the project by building the organisational structure, fostering local and international partnerships, and setting up the collaborative frameworks necessary for successful data integration and project implementation. This foundational work ensures that subsequent research activities are well-coordinated, scientifically robust, and closely aligned with stakeholder needs.

#### WP2 – Ph.D. project “Social health inequalities in the first 1000 days of life in Kalaallit Nunaat”

WP2 is designed as a Ph.D. project aimed at investigating health and health inequalities among children aged 0-2 in Kalaallit Nunaat. It will explore how parental socioeconomic status, influence health outcomes and investigate how early prevention programs affect child health outcomes. This work package primarily focuses on data from the healthcare system to identify key determinants of child health and work with local health care practitioners to validate and discuss registration practices for small children’s health.

The healthcare system represents potentially the largest and most validated data source concerning children’s health and well-being, with a vast and complex dataset that necessitates dedicated and specialized attention. To address this, a PhD project has been established specifically to manage, analyse, and interpret these data through a social inequality lens. This work package is a direct extension of a previous and published pilot study and provides a focused investigation of healthcare system data (39). The insights gained will inform policies on early intervention and public health strategies aimed at reducing disparities and improving health outcomes among vulnerable child populations.

#### WP3 – Post. doc project: “Effects of determinants and ACEs on children’s health - investigating health outcomes and trajectories for children aged 2-18”

WP3 is designed as a post doc study, to investigate the complex effects of social determinants and ACEs on the health of children and adolescents aged 2 to 18 years in Kalaallit Nunaat. This work package aims to characterize the health status of this population; identify key social determinants and ACEs they face as well as identifying health promoting factors for child health and well-being. The project further aims to analyze how clusters of social determinants during the first 1000 days of life influence health trajectories throughout childhood and adolescence and explores the dynamic associations between trajectories of ACEs and health outcomes over time, providing insight into complex life course mechanisms.

WP3 works broadly across multiple data sources that have never before been combined or used for research and monitoring, integrating information from various registers to provide a comprehensive, cross-sectoral perspective. Employing advanced methods such as latent class analysis (LCA) and trajectory analyses, WP3 offers a life-course approach to understanding health inequalities. The findings will complement WP2 and contribute to the overall project by informing targeted multi-dimensional strategies for health promotion as well as prevention and intervention addressing inequities in health and well-being.

### Expected outcomes

The project will establish proof-of-concept for a monitoring system that in the continuation will provide fact based and always updated knowledge on children’s health in years to come. The proof-of-concept aims to harvest existing data sources, often unstructured, collect and disseminate statistical tables to support knowledge sharing in already established systems as well as allowing researchers to access collected data in combination with other data held by Statistics Greenland. Additionally, one of the expected outcomes of the project is to produce an annual report on the health and wellbeing of all children in Kalaallit Nunaat ensuring continuous and accessible data to support children’s health. The report will be integrated into the systems of Statistics Greenland, which will ensure that annual reporting continues beyond the project period.

### Ethics

The study was approved by the Scientific Ethics Committee in Kalaallit Nunaat (VEK 2025-12), and the SDU Research and Innovation Organization (SDU RIO), in accordance with the GDPR (EU) 2016/679. The study aligns with the Kalaallit Nunaat government’s research strategy for 2022-2030 (46) as the study aims to supporting sustainable societal development by contributing to the improvement of public health and the reduction of social health inequalities.

### Dissemination of results

The project results will be disseminated through a mix of scientific publications, practitioner engagement, and public communication. A total of seven peer-reviewed articles are planned: three from WP2 and four from WP3. In addition, an annual child health statistic published from Statistics Greenland will be produced to support national monitoring and inform the political and administrative level about children’s health and well-being.

A key part of the dissemination strategy is regular meetings with local practitioners, where findings will be presented through dialogue in ways that are meaningful for practice and support reflection on data use. This dissemination strategy builds on the close and respectful relationships that are formed between the project group and key stakeholders for children’s health. The project also aims to reach a broader audience through popular science platforms and social media. Two introductory articles have already been published online in collaboration with midwifes and community health nurses to help make results accessible to the general population (47, 48).

## Discussion

This study describes the protocol for the research project Meeqqat Peqqissut, which aims to strengthen child and adolescent health monitoring in Kalaallit Nunaat through a cross-sectoral, register-based approach. This study provides an important contribution to public health research in Kalaallit Nunaat by establishing a comprehensive and sustainable register-based infrastructure that includes all children in the country and enabling longitudinal follow-up from birth through adolescence across the different sectors. This novel approach provides a unique opportunity to fill a substantial knowledge gap regarding children’s health and well-being.

This study marks an important first step toward building similar capacities in Kalaallit Nunaat as have already been put in place in countries like Norway and Denmark, where longitudinal infrastructures such as the SamBa study and the Child Health Database are well-established. By initiating a national, cross-sectoral register-based platform, the project addresses the current lack of systems to systematically track children’s health and well-being over time. In Denmark, infrastructures such as the Child Health Database have enabled municipalities to develop cohort-specific health profiles based on community health nurse data and use them for both public health planning and monitoring (23, 49). The Child Health Database has also made the possible to provide longitudinal data on early child health indicators and later diagnosis of mental disorders (50–52). The DANLIFE cohort further demonstrates how systematically collected register data can be used to study the long-term effects of ACEs on health trajectories (25, 53).

While this project draws inspiration from these databases, it also distinguishes itself through its focus on Indigenous Arctic contexts and its strong emphasis on community engagement. Moreover, this study navigates unique challenges related to fragmented data systems, evolving electronic health records, and the integration of health promoting factors and culturally relevant indicators. In doing so, it offers not just a technical solution, but a model for how longitudinal, equity-focused child health surveillance can be built from the ground up in a culturally distinct setting.

### Strengths and limitations

The inclusion of the entire population in the study strengthens the validity and generalizability of findings and allows for detailed examination of life course processes, which is crucial for identifying critical windows of intervention and health promotion. A key strength is the cross-sectoral integration of data from health care, municipalities, Statistics Greenland, and the police, which creates a uniquely holistic perspective on children’s lives, capturing both their health and the social determinants shaping their well-being. This enables the identification of complex patterns and interactions across sectors that are rarely accessible in traditional health research.

The participatory approach, grounded in community-based participatory research principles, further strengthens the project by involving practitioners and other local stakeholders throughout the research process. This ensures that the data is not only technically valid but also meaningful and actionable within the local context, and it promotes shared responsibility for data quality and relevance. Another innovative contribution lies in the project’s attempt to operationalize strength-based approaches within register-based research. While administrative data typically focuses on problems such as illness, service use, or adverse events, this study actively seeks to identify and incorporate health promoting factors and culturally relevant indicators.

However, the study also faces several limitations that need to be considered. First, register data is only as reliable as the systems and practices used to generate them. In Kalaallit Nunaat, variation in registration practices across institutions, regions, and over time can affect data quality and comparability. This makes it essential to complement data analysis with qualitative insight into registration practices, which the project addresses by engaging practitioners in discussions on data validity and contributing to capacity-building in data documentation and standardization.

Second, technical limitations related to the use of various electronic patient journal (EPJ) systems, such as COSMIC, Æskulap, and Get Organized create challenges for data accessibility and continuity. The phased implementation of systems and uneven uptake across regions and municipalities, means that national coverage is incomplete for some years. Moreover, the upcoming replacement of COSMIC introduces uncertainty about future data structures and access. Following the end of the Greenlandic-Danish collaboration project period in 2023, the Agency of Social Affairs has experienced a reduction in available resources, which has limited its capacity to monitor and support the ongoing implementation of initiatives. The consequence for this project is at this point unknown.

Third, historical data are often difficult to validate and may be incomplete, which limits the ability to conduct robust retrospective analyses. Consequently, the project primarily focuses on developing a prospective, continuously updated data infrastructure to support future monitoring and longitudinal research. While this forward-looking approach strengthens the infrastructure for long-term surveillance, it does constrain the ability to examine historical trends in depth. Some historical data are available through Statistics Greenland, and the project team will make use of all accessible sources while carefully ensuring that only reliable and validated measures are included in the analyses.

Lastly, while the project seeks to incorporate a strength-based perspective. This necessitates careful methodological work to pinpoint the specific health promoting factors on different levels that might be recorded in the systems including the qualitative registrations and free writing in the registration systems.

### Public health implications

This project contributes directly to strengthening the public health infrastructure in Kalaallit Nunaat by enabling more systematic use of existing data across sectors. Through the integration of cross sectoral data, it becomes possible to identify patterns in children’s health and well-being, and to understand how determinants in terms of both risk- and health promoting factors, affects health outcomes over time.

The project supports the implementation of the current public health program, *Inuuneritta III*, which focuses on providing a good and safe life for all children. The program calls for a unified and regularly updated child health database to monitor and guide public health efforts. By consolidating data through Statistics Greenland, the project establishes a foundation for monitoring.

## Conclusion

The project supports the national public health programme *Inuuneritta III* by contributing to the development of a unified and validated regularly updated child health data infrastructure combining data that has never been combined or used for research before.

By integrating data from multiple sectors and anchoring the work in local collaboration and strength-based perspectives, *Meeqqat Peqqissut* offers a more holistic understanding of social health inequalities in an Indigenous Arctic context. Despite limitations in data quality and system infrastructure, the project lays the groundwork for a sustainable and scientifically robust monitoring system. It represents a critical step toward earlier identification of health and social risks as well as health promoting factors, more targeted public health efforts, and the long-term goal of reducing social inequalities in health among children and youth in Kalaallit Nunaat.

## Data Availability

For Study Protocols: No datasets were generated or analysed during the current study. All relevant data from this study will be made available upon study completion.

## Acknowledgements

We would like to thank all members of the project’s reference group for their contributions to the project definition. Furthermore, we are grateful to all participants in the scientific advisory board for their valuable feedback on this article. We thank Trine P. Pedersen (University of Southern Denmark), Lau C. Thygesen (University of Southern Denmark), and Marin Strøm (University of the Faroe Islands) for taking the time to review the manuscript.

## References

1. Niclasen B, Köhler L. National indicators of child health and well-being in Greenland. Scand J Public Health. 2009;37(4):347–56.

2. Albertsen N, Lynge AR, Skovgaard N, Olesen JS, Pedersen ML. Coverage rates of the children vaccination programme in Greenland. Int J Circumpolar Health. 2020;79(1):1721983.

3. Børresen ML, Koch A, Biggar RJ, Ladefoged K, Melbye M, Wohlfahrt J, Krause TG. Effectiveness of the targeted hepatitis B vaccination program in Greenland. Am J Public Health. 2012;102(2):277–84.

4. Ekstrand KR, Abreu-Placeres N. The impact of a national caries strategy in Greenland 10 years after implementation. A failure or a success? Int J Circumpolar Health. 2020;79(1):1804260.

5. Friborg J, Koch A, Stenz F, Wohlfahrt J, Melbye M. A population-based registry study of infant mortality in the Arctic: Greenland and Denmark, 1973-1997. Am J Public Health. 2004;94(3):452–7.

6. Hansen CH, Koch A, Wohlfahrt J, Melbye M. A population-based register study of vaccine coverage among children in Greenland. Vaccine. 2003;21(15):1704–9.

7. Jespersen SI, Demant MN, Pedersen ML, Homøe P. Acute otitis media and pneumococcal vaccination - an observational cross-sectional study of otitis media among vaccinated and unvaccinated children in Greenland. Int J Circumpolar Health. 2021;80(1):1858615.

8. Tindborg M, Koch A, Andersson M, Juul K, Geisler UW, Soborg B, Michelsen SW. Heart disease among Greenlandic children and young adults: a nationwide cohort study. Int J Epidemiol. 2022;51(5):1568–80.

9. Christensen LH, Høyer BB, Pedersen HS, Zinchuk A, Jönsson BAG, Lindh C, et al. Prenatal smoking exposure, measured as maternal serum cotinine, and children’s motor developmental milestones and motor function: A follow-up study. Neurotoxicology. 2016;53:236–45.

10. Hahn GH, Koch A, Melbye M, Mølbak K. Pattern of drug prescription for children under the age of four years in a population in Greenland. Acta Paediatr. 2005;94(1):99–106.

11. Kløvgaard M, Nielsen NO, Sørensen TL, Bjerregaard P, Olsen B, Christesen HT. Children in Greenland: disease patterns and contacts to the health care system. Int J Circumpolar Health. 2016;75:32903.

12. Kløvgaard M, Nielsen NO, Sørensen TL, Bjerregaard P, Olsen B, Júlíusson PB, et al. Growth of children in Greenland exceeds the World Health Organization growth charts. Acta Paediatr. 2018;107(11):1953–65.

13. Kok Grouleff M, Wielsøe M, Berthelsen D, Mulvad G, Isidor S, Long M, Bonefeld-Jørgensen EC. Anthropometric measures and blood pressure of Greenlandic preschool children. Int J Circumpolar Health. 2021;80(1):1954382.

14. Ottendahl CB, Bjerregaard P, Svartá DL, Seidler IK, Olesen I, Nielsen MS, Larsen CVL. Childhood conditions and mental health among youth and young adults in Greenland: a latent class analysis. International Journal of Circumpolar Health. 2024;83(1):2400397.

15. Merrick MT, Ford DC, Ports KA, Guinn AS, Chen J, Klevens J, et al. Vital Signs: Estimated Proportion of Adult Health Problems Attributable to Adverse Childhood Experiences and Implications for Prevention - 25 States, 2015-2017. MMWR Morb Mortal Wkly Rep. 2019;68(44):999–1005.

16. Ottendahl CB, Seidler IK, Beck A, Pedersen CP, Bjerregaard P, Larsen CVL. Developing the ACEIG-scale: An adverse childhood experience scale for Inuit youth in Greenland. Child Abuse Negl. 2024;148:106471.

17. Seidler IK, Tolstrup JS, Bjerregaard P, Crawford A, Larsen C. Time trends and geographical patterns in suicide among Greenland Inuit. BMC Psychiatry. 2023;23(1):187.

18. The Greenlandic Health Commission. Sundhedskommissionens betænkning - Vores sundhedsvæsen - Vores fælles ansvar [The Health Commission’s report - Our health care system - our joint responsebility, in Danish]. Nuuk; 2023.

19. Naalakkersuisut. INUUNERITTA III - Naalakkersuisuts strategi for samarbejdet om det gode børneliv 2020-2030 [The Government of Greenlands strategy for the collaboration for creating good lives for all children, in Danish] Nuuk: The Department of social affairs, Families and Law, The Depatment of Education, Culture and Church, The Department of Health 2020.

20. Ingemann C, Kvernmo S, Møller H, Moffitt PM, Tagalik S, Kuhn RL, et al. Symposium on “parental education” at the ICCH17. Int J Circumpolar Health. 2019;78(1):1604062.

21. Hansen KL, Fluke J, Gesink D, Friborg O, Sørlie T, Merkel-Holguin L, Martinussen M. Study Protocol: The Arctic Childhood Study: a Study of Violence and Health in Indigenous Sámi and Non-Sámi Children and Youth in Arctic Norway—a Mixed Methods Cohort Study Design. International Journal on Child Maltreatment: Research, Policy and Practice. 2023.

22. National Institute of Public Health. Rapporter baseret på data fra Databasen Børns Sundhed [Reports based on The Database for Childresn Health, in Danish] 2023 [Available from: https://www.sdu.dk/da/sif/forskning/projekter/databasen_boerns_sundhed/publikationer/rapporter.

23. Pommerencke LM, Pant SW, Madsen KR, Laursen B, Pedersen TP. Social ulighed i børn og unges udvikling, sundhed og trivsel [Social inequalities in children’s and youth’s development, health and well-being, in Danish]. Copenhagen National Institute of Public Health 2022.

24. Quality institute in the Danish Healthcare system. Landsdækkende Database for Børn og Unges Sundhed [National data base for children and adolescent health. In Danish] 2025 [Available from: https://www.sundk.dk/kliniske-kvalitetsdatabaser/landsdaekkende-database-for-boern-og-unges-sundhed-ldbu/.

25. Bengtsson J, Dich N, Rieckmann A, Hulvej Rod N. Cohort profile: the DANish LIFE course (DANLIFE) cohort, a prospective register-based cohort of all children born in Denmark since 1980. BMJ Open. 2019;9(9):e027217.

26. Marmot M. Social determinants of health inequalities. The Lancet. 2005;365(9464):1099–104.

27. Bronfenbrenner U. Ecological models of human development. International encyclopedia of education. 1994;3(2):37–43.

28. Fish J, Hirsch G, Syed M. “Walking in Two Worlds”: Toward an Indigenist Ecological Systems Model for Group Therapy. The Counseling Psychologist. 2022;50(5):622–48.

29. O’Keefe VM, Fish J, Maudrie TL, Hunter AM, Tai Rakena HG, Ullrich JS, et al. Centering Indigenous Knowledges and Worldviews: Applying the Indigenist Ecological Systems Model to Youth Mental Health and Wellness Research and Programs. Int J Environ Res Public Health. 2022;19(10).

30. Guy-Evans O. Bronfenbrenner’s Ecological Systems Theory 2024 [Available from: https://www.simplypsychology.org/bronfenbrenner.html.

31. Olesen I, Larsen CVL. Peqqissuserput: A theoretical research model starting from a place of strengths to support Kalaallit Inuit communities to thrive. Journal of Community Systems for Health. 2025;2(2).

32. Thygesen LC, Ersbøll AK. When the entire population is the sample: strengths and limitations in register-based epidemiology. European Journal of Epidemiology. 2014;29(8):551–8.

33. Juul S. Epidemiologi og evidens [Epidemiology and evidence, in Danish]. 3. ed ed. Kbh.: Munksgaard; 2017. 328 pages p.

34. Statistics Greenland. Befolkningen pr 1. januar 1977-2025 [Population 1. January 1977-2025. In Danish] 2025 [Available from: https://bank.stat.gl/pxweb/da/Greenland/Greenland__BE__BE01__BE0120/BEXSTA.px/.

35. Pedersen CB. The Danish Civil Registration System. Scand J Public Health. 2011;39(7 Suppl):22–5.

36. CAMBIO. Cambio COSMIC health care information system 2025 [Available from: https://www.cambiogroup.com/our-solutions/cambio-cosmic/?utm_source=chatgpt.com.

37. Naalakkersuisut. Svar § 37 spørgsmål nr. 163-2024 [Answer § 37 question nr. 163-2024] 2024 [Available from: https://ina.gl/media/v4nnsl5p/163_2024_patientjournalsystem_mettah_svar.pdf.

38. National Institute of Public Health SDU. Social inequality in health among 0-1 year olds in Greenland 2023 [cited 2023. Available from: https://www.sdu.dk/en/sif/forskning/projekter/social_ulighed_blandt_0_1_aarige_i_groenland.

39. Ottendahl CB, Sivholm EA, Ritz C, Hansen KL, Larsen CVL. Social inequalities in health among children born in 2022 in Kalaallit Nunaat - A national register-based study with insights from local healthcare practitioners. Int J Circumpolar Health. forthcoming.

40. Agency of Social Affairs, Department for Children Youth and Families. SAMARBEJDE FOR BØRN OG UNGE - Grønlandsk-dansk tværgående arbejde [Collaboration for children and youth - Greenlandic-Danish crosssectional collaboration, in Danish]. 2024.

41. Naalakkersuisut. Inatsisartutlov om ændring af Inatsisartutlov om støtte til børn 2023 [Available from: https://nalunaarutit.gl/groenlandsk-lovgivning/2023/inatsisartutlov-nr-68-af-20_11_2023?sc_lang=da.

42. Naalakkersuisut. Grønlandsk-dansk tværgående arbejde for en styrket indsats for udsatte børn og unge i Grønland, 2020-2023 - Afsluttende rapport [Greenlandic-Danish Cross-Sectoral Collaboration for Strengthened Efforts for Vulnerable Children and Youth in Greenland, 2020-2023 - Final Report, in Danish]. 2024.

43. Minkler M, Wallerstein N. Community-based participatory research for health: From process to outcomes: John Wiley & Sons; 2011.

44. Peterson M, Augustine R, Elizabeth R, Mark S, Julia H, Gitte AR, and Larsen CV. Applying community-based participatory research principles to build trust and equity in health and socio-ecological studies in Greenland. International Journal of Circumpolar Health. 2025;84(1):2473181.

45. Cueva K, Rink E, Lavoie JG, Stoor JPA, Healey Akearok G, Gladun E, Larsen CVL. Diving below the surface: A framework for arctic health research to support thriving communities. Scandinavian Journal of Public Health. 2021.

46. Naalakkersuisut. Ilisimatusarneq – ineriartornermut aqqutissaavoq Kalaallit Nunaanni ilisimatusarnermut periusissiaq [Research - the road a head. Greenland’s national research startegy. In Greenlandic]. 2022.

47. Ottendahl CB, Willesen A, Berthelsen D, Larsen CVL. Sådan kan forskning forbedre grønlandske børns liv [This is how research kan improve child health in Kalaallit Nunaat. In Danish] [Online newspaper]. 2025 [Available from: https://videnskab.dk/krop-sundhed/saadan-kan-forskning-forbedre-groenlandske-boerns-liv/.

48. Ottendahl CB, Willesen A, Berthelsen D, Larsen CVL. Små børns sundhed i Grønland: Hvad ved vi, og hvad mangler vi fortsat svar på? [Small chiildrenøs health in Kalaallit Nunaat. What do we know and where do we still need answers? In Danish] [Online newspaper]. 2025 [Available from: https://www.sundhedsplejersken.nu/smaa-boerns-sundhed-i-groenland-hvad-ved-vi-og-hvad-mangler-vi-fortsat-svar-paa/.

49. Jørgensen SE, Pommerencke LM, R CR, Pedersen TP. Sundhedsprofil for børn og unge i Region Hovedstaden og kommuner - Baseret på sundhedsplejerskedata om børn født i 2021 og børn og unge ind- og udskolingsundersøgt i 2021/22 [Health profile for children and adolescents in the capital region and municipalities – based on health visitor data on children born in 2021 and children and adolescents examined at school entry and exit in 2021/22. In Danish]. Statens Institut for Folkesundhed SDU, København: Statens Institut for Folkesundhed SDU; 2023.

50. Pant SW, Holstein BE, Ammitzbøll J, Skovgaard AM, Pedersen TP. Community health nurses’ concerns about infant regulatory problems are predictive of mental disorders diagnosed at hospital: a prospective cohort study. Front Child Adolesc Psychiatry. 2023;2:1330277.

51. Pant SW, Skovgaard AM, Ammitzbøll J, Holstein BE, Pedersen TP. Motor development problems in infancy predict mental disorders in childhood: a longitudinal cohort study. Eur J Pediatr. 2022;181(7):2655–61.

52. Holstein BE, Pant SW, Ammitzbøll J, Laursen B, Madsen KR, Skovgaard AM, Pedersen TP. Parental education, parent-child relations and diagnosed mental disorders in childhood: prospective child cohort study. Eur J Public Health. 2021;31(3):514–20.

53. Rod NH, Bengtsson J, Budtz-Jorgensen E, Clipet-Jensen C, Taylor-Robinson D, Andersen AN, et al. Trajectories of childhood adversity and mortality in early adulthood: a population-based cohort study. Lancet. 2020;396(10249):489–97.

